# The impact of climate variability on dengue virus case incidence in Sindh, Pakistan: Post-2022 flooding

**DOI:** 10.64898/2026.11.05.26367382

**Authors:** Amna Tariq, Raheel Allana, Sajid Shah, Momin Kazi, Desiree LaBeaud

## Abstract

Dengue virus is spread primarily by the Aedes aegypti and Ae. albopictus mosquitoes is a self-limiting viral disease primarily present in the tropical and subtropical regions of the world. Temperature and rainfall impact vector development, virus replication, and habitat suitability for Ae. aegypti and Ae. albopictus larvae and pupae. Pakistan, located in Southeast Asia, is one country where DENV is endemic and impacts thousands of lives each year. In recent years, Pakistan has been hardly hit by climate change, with heavy rainfalls and flooding recurring each year, with simultaneous surges of dengue virus across the country. In this paper, we analyze the monthly dengue virus disease data from Sindh Pakistan, from 2020-2025 to understand the impacts of flooding, rainfall, and temperature fluctuations on dengue case incidence in the region. We use parametric generalized linear models and the generalized additive model. Our results indicate that monthly confirmed dengue cases exhibit a strong seasonality, with the great majority of cases concentrated in the post-monsoon months (September–November). A 1-month lag in rainfall was associated with a statistically significant increase in dengue incidence (IRR = 1.008, 95% CI: 1.003–1.012, p = 0.002), corresponding to a 0.8% increase in expected case counts per unit (mm) increase in rainfall one month prior. In contrast, neither lagged temperature term reached statistical significance.

## Introduction

Dengue virus (DENV) disease is a self-limiting arthropod borne viral infection primarily reported in the tropical and subtropical regions of the world where conditions of temperature and humidity favor the proliferation of its mosquito vectors [1-3]. DENV is a *flavivirus* and is horizontally spread through the bite of female *Aedes aegypti* and *Aedes Albopictus* mosquitoes [3-5]. There are four distinct antigenic circulating serotypes of DENV, namely DENV-1, DENV-2, DENV-3, and DENV-4 [6]. The disease usually presents as fever, headache, vomiting, muscle pain, joint pain, and a distinctive itching and skin rash that usually begins 3–4 days after infection [5, 7]. DENV has been reported in more than 100 tropical and sub-tropical countries around the globe [1]. Each year, approximately 400 million cases and 22,000 deaths occur due to DENV [1]. Studies have reported approximately 2.5 billion people at risk of dengue transmission and about 50 million new dengue infections each year [8]. Due to a high global DENV burden and climate change as an imminent threat to human population, recent studies have been more focused on understanding the impacts of climatic parameters such as precipitation, humidity and temperature on the spread of DENV [9].

Climate change impacts DENV and Aedes vector population, both directly and indirectly [10]. Temperature variation influences vector development rates, mortality, and behavior and impacts virus replication within mosquitoes [11-13]. Precipitation impacts habitat suitability for *Ae. aegypti* and *Ae. albopictus* larvae and pupae. Temperature also interacts with rainfall regulating evaporation, thereby also affecting the availability of water habitats for mosquitoes[9]. Indirectly, temperature, rainfall, and humidity influence land cover and use, which can accelerate or impede the growth of vector populations [9].

Pakistan, located in Southeast Asia, is one country where DENV is endemic and impacts thousands of lives each year [14]. With a population of over 210 million, DENV has emerged as a major public health concern [14, 15]. In recent years, Pakistan has also been the worst recipient of climate change, with heavy rainfalls and flooding recurring each year [16]. In 2022 alone, the country received rainfall equivalent to 2·9 times its national 30-year average [16]. These heavy rainfalls create a suitable breeding environment for *Aedes aegypti* mosquitoes, resulting in surges of dengue virus each year [16]. In Pakistan, climate change, along with globalization, demographic changes, trade, and global warming, are mainly responsible for the spread of *Aedes aegypti* vectors by creating environments suitable for mosquito proliferation [17].

Pakistan reported the first ever presence of DENV in 1994, and the number of laboratory-confirmed cases has steadily increased since then. Pakistan has reported notable DENV outbreaks in 2010, 2017, 2019, 2020, and 2021 [18]. During the most recent massive DENV outbreak in Pakistan in 2022, Karachi, a coastal city located in the province of Sindh, was among the cities with the highest number

of reported DENV cases [19, 20]. During the same year, Pakistan was hit by unusually heavy torrential monsoon rains and the province of Sindh was affected the most [21]. As the region continues to experience recurring heavy rains and continued substantial DENV burden, the epidemiological relationship between rainfall, temperature, humidity and dengue case incidence needs to be adequately characterized. In this manuscript, we analyze the monthly dengue virus disease data from Sindh Pakistan to understand the impacts of flooding, rainfalls and temperature fluctuations on case incidence.

### Ethics statement

The study protocol was approved by the Stanford University Institutional Review Board (Protocol ID #31488) and the Ethical Review Board at Agha Khan Univerisity (protocol ID#).

## Methods

### Data

Province specific monthly dengue virus confirmed case data from 2020-2025 was obtained from the Sindh ministry of Health, Pakistan. We retrieved province specific monthly mean, maximum and minimum values on precipitation, humidity and temperature publicly available from Visual Crossings from 2020-2025 [22]. The monthly mean value was computed using province-specific daily temperature observations and the monthly aggregated rainfall was computed using province-specific daily rainfall data.

### Statistical Analysis

We fit and compared the parametric generalized linear models (GLMs) and the generalized additive model (GAM) to the dengue case incidence data. Since the dengue case counts were overdispersed (variance exceeds the mean), therefore, a Negative Binomial (NB) error distribution was used throughout rather than Poisson, with a log link. A cross-correlation relationship between temperature and DENV cases and rainfall and DENV cases was assessed. Since literature states that dengue transmission depends on mosquito breeding and an extrinsic incubation period rather than same-month weather, we used lagged rainfall and temperature before modeling. Rainfall was utilized at 1- and 2-month lags (reflecting the time needed for standing water to accumulate and larvae to mature), and temperature was utilized at 2- and 3-month lags (reflecting the incubation period linking warmer weather to detectable case increases). This is consistent with lag structures reported in the dengue-climate literature and those derived from our cross correlations. Observations with missing lagged values (the first 3 months of the series) were dropped, leaving n = 65 for model fitting.

For the parametric GLM, we tested several seasonal specifications (a) categorical month dummies, (b) first harmonic (annual sine/cosine) seasonality, (c) first + second harmonic seasonality, and variants substituting log-transformed rainfall, cumulative 2-month rainfall, and a linear time trend. All GLM variants included the same rainfall and temperature lag terms as linear predictors. For the GAM, we allowed each predictor a flexible, non-linear shape rather than assuming a fixed log-linear effect: a cyclic penalized spline for calendar month (to enforce December–January continuity), penalized splines (k = 4 basis functions, given the limited sample size) for each rainfall and temperature lag, and a linear time trend.

Candidate GLM specifications were first screened on in-sample AIC/BIC and pseudo-R^2^ (1 − deviance/null deviance). However, because in-sample fit statistics reward overfitting — particularly for the 16-parameter month-dummy model relative to only 65 observations — the primary selection criterion was rolling-origin (walk-forward) cross-validation: starting from a 30-month training window, each candidate model was refit and used to forecast only the single next unseen month, the window was expanded by one month, and this was repeated to the end of the series (35 out-of-sample forecasts per model). Models were ranked by root mean squared error (RMSE) across these forecasts, which approximates real forecasting performance rather than retrospective fit. The same rolling-origin procedure was used to compare the best-performing GLM against the GAM.

All statistical analysis were conducted in R, Version 2026.01.1+403 (2026.01.1+403)

### Results

Over the study period (January 2020–August 2025, n = 68 months), monthly confirmed dengue cases exhibited strong seasonality, with the great majority of cases concentrated in the post-monsoon months (September–November) and near-zero counts for much of the rest of the year. A single extreme outbreak dominated the series: monthly cases rose from a baseline of under 300 to a peak of 8,935 in September 2022, coinciding with anomalously heavy monsoon rainfall that year. Excluding this outbreak, seasonal peaks in other years ranged from approximately 600 to 2,100 cases per month. From 2017-2022, 61,853 DENV cases were lab confirmed from the 45,1527 DENV screened at the primary and tertiary care hospitals. The highest number of cases 23754 were reported in 2022 with 62 deaths. The mean temperature over the six years averaged to 80.8°F (SD: 14.2) and the cumulative monthly rainfall averaged at 15.8mm (40.2). The mean DENV cases reported each month were 593.6 (SD: 1457.1).

**Figure 1.**
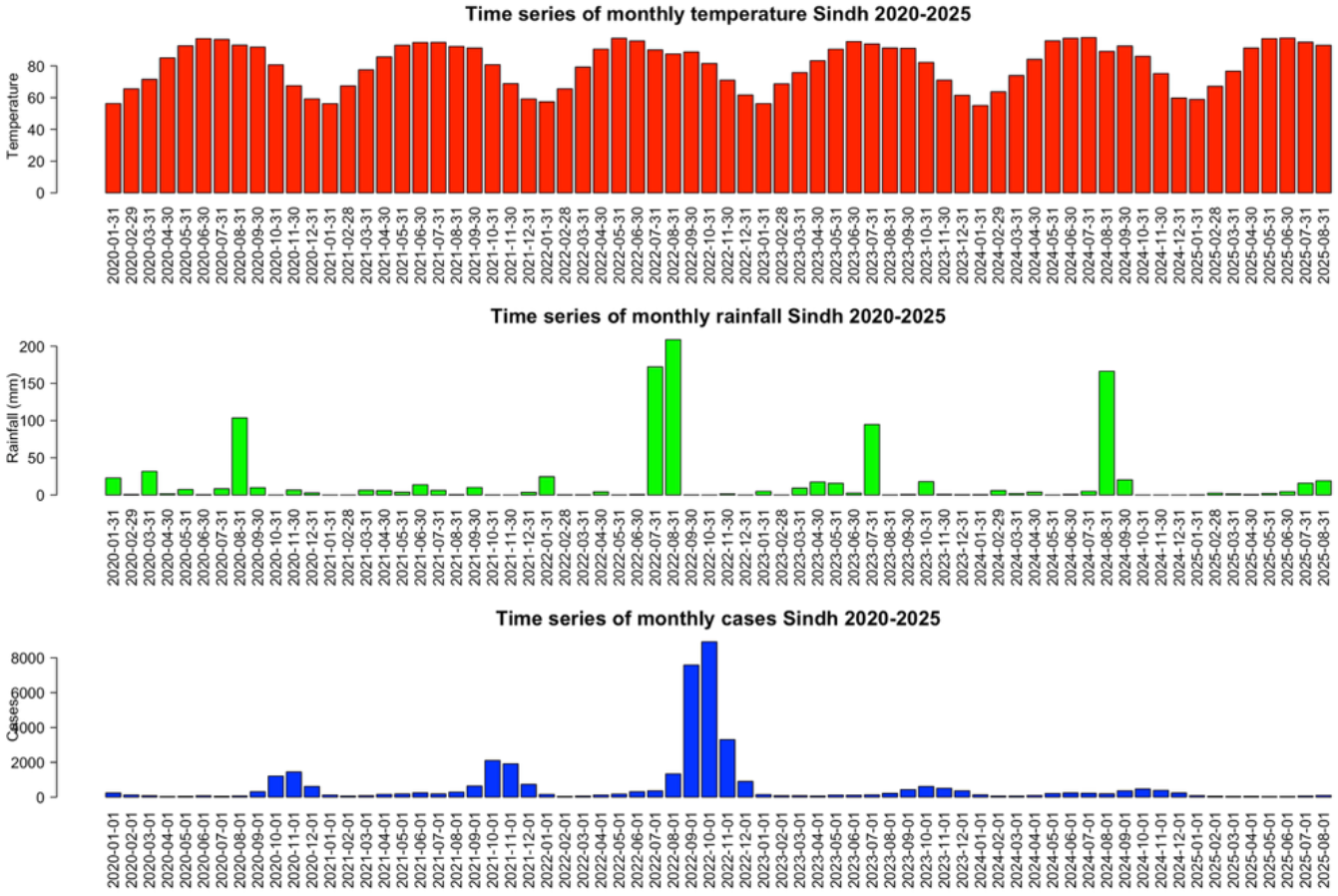
Panel plot showing the time series of monthly temperature rainfall and DENV cases from 2020-2025.

**Figure 2.**
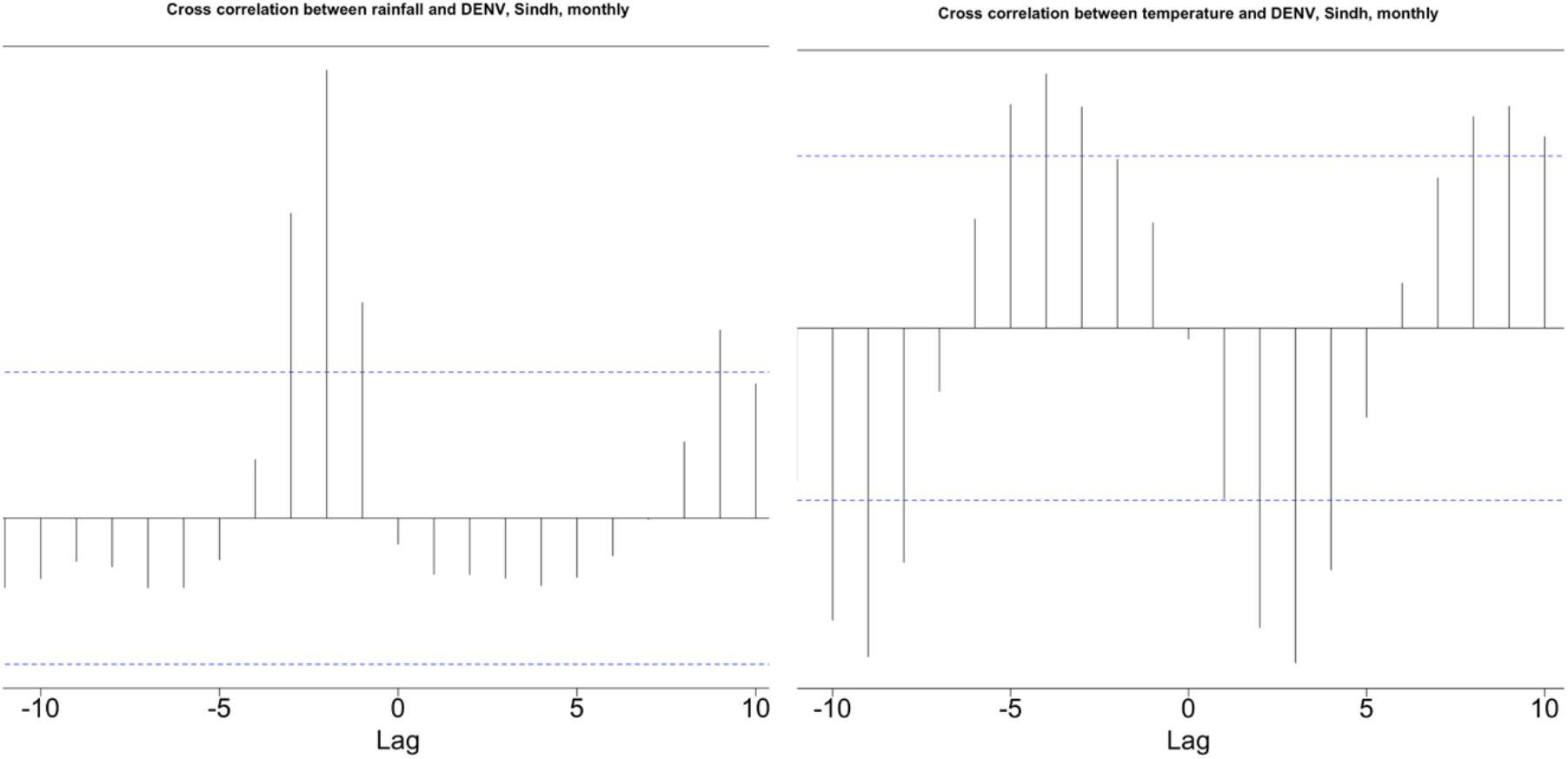
Cross correlation function of DENV cases with rainfall and temperature in Sindh Pakistan.

The negative binomial GLM identified rainfall, but not temperature, as a significant predictor of monthly dengue case counts after adjusting for seasonality. The negative binomial GLM model explained a substantial proportion of deviance in monthly dengue case counts, with a deviance-based pseudo-R^2^ of 0.830. The estimated dispersion parameter (θ = 2.868) confirmed meaningful overdispersion in the case count data relative to a Poisson model, supporting the use of a negative binomial specification.

A 1-month lag in rainfall was associated with a statistically significant increase in dengue incidence (IRR = 1.008, 95% CI: 1.003–1.012, p = 0.002), corresponding to a 0.8% increase in expected case counts per unit (mm) increase in rainfall one month prior. A 2-month rainfall lag showed a similar, slightly smaller effect (IRR = 1.006, 95% CI: 1.001–1.011, p = 0.027), indicating that elevated rainfall continues to predict higher case counts up to two months later, consistent with the time needed for standing water to support mosquito breeding and larval development.

In contrast, neither lagged temperature term reached statistical significance. Temperature lagged by 2 months showed a coefficient in the opposite direction from expectation (IRR = 0.965, 95% CI: 0.874– 1.067, p = 0.436), and temperature lagged by 3 months showed a near-null effect (IRR = 0.986, 95% CI: 0.902–1.078, p = 0.736). Both confidence intervals span 1, indicating no evidence that temperature, at these lags, independently predicts dengue case counts once rainfall and (if included) seasonal terms are accounted for.

Overall, these results suggest rainfall, particularly at short lags of 1–2 months is the more robust climate driver of dengue transmission in this dataset, while the temperature signal is weak or absent at the lags tested.

**Figure 3.**
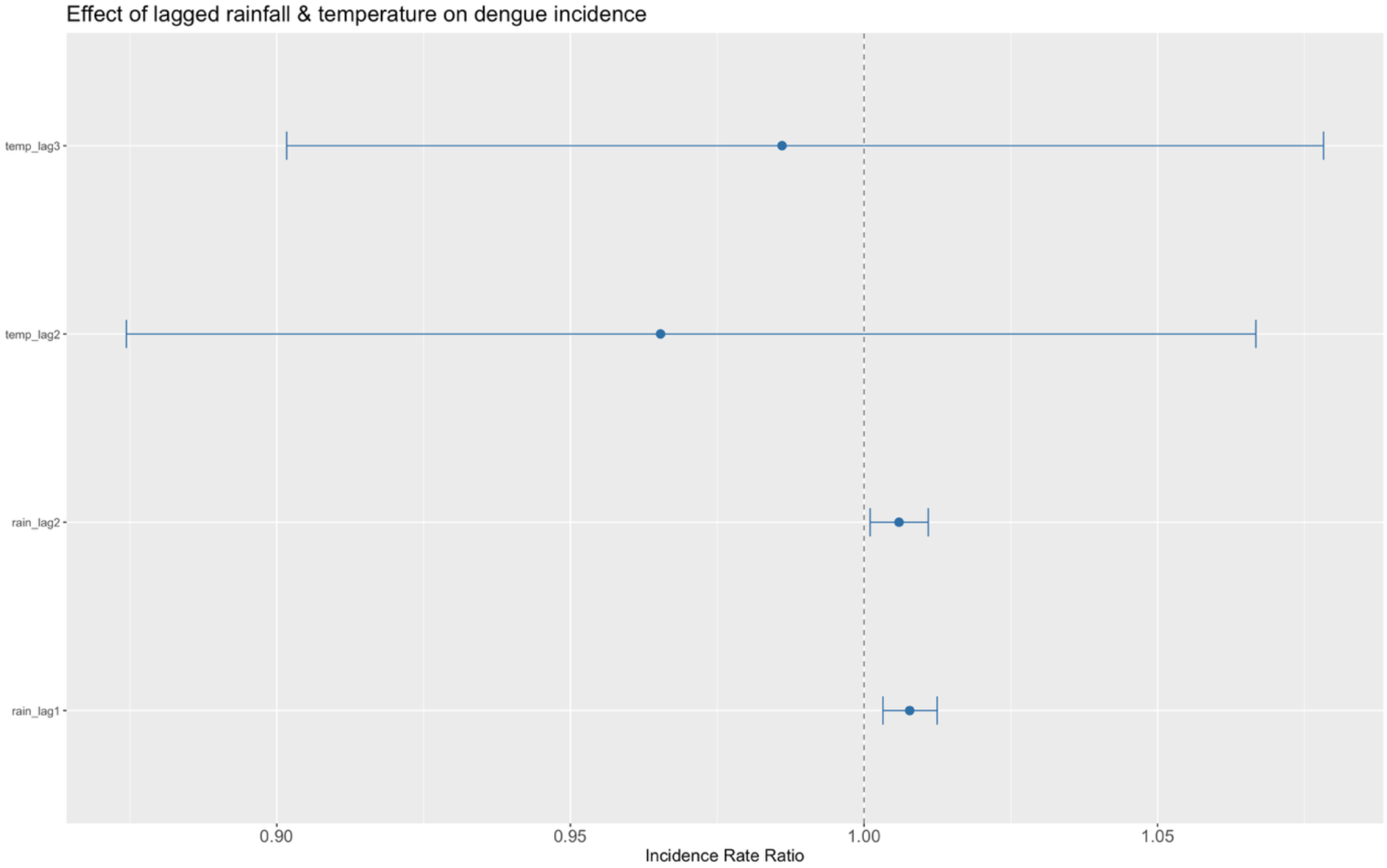
Effect of lagged temperature and rainfall on dengue case incidence

### Model comparison

Seven candidate Negative Binomial generalized linear model (GLM) specifications, differing in their representation of seasonality (categorical month indicators vs. first- and second-order harmonic terms) and rainfall parameterization (raw, log-transformed, or cumulative), were compared using rolling-origin cross-validation (30-month initial training window, 35 sequential one-step-ahead forecasts).

The best-performing specification combined a first- and second-harmonic seasonal term, a linear time trend, and linear terms for rainfall (1- and 2-month lag) and temperature (2- and 3-month lag), achieving the lowest out-of-sample RMSE (1,192.9) among all candidates (Table 1). Notably, the fully saturated categorical-month specification, despite comparable in-sample pseudo-R^2^ (0.83 vs. 0.833), produced the second-worst out-of-sample RMSE (1,752.4), consistent with overfitting given the limited sample size relative to its 16 estimated parameters.

**Table 1.** Out-of-sample performance of candidate GLM specifications (ranked by RMSE)

| **Model** | **Parameters** | **Pseudo R<sup>2</sup>** | **AIC** | **Out-of-sample RMSE** |
|---|---|---|---|---|
| Harmonic seasonality + trend | 10 | 0.833 | 838.5 | **1,192.9** |
| Harmonic seasonality (1st order) | 7 | 0.782 | 850.8 | 1,307.8 |
| Harmonic + cumulative rainfall | 8 | 0.823 | 838.4 | 1,429.6 |
| Harmonic, rainfall only | 7 | 0.819 | 838.1 | 1,431.2 |
| Harmonic seasonality (1st + 2nd order) | 9 | 0.824 | 840.2 | 1,559.4 |
| Categorical month indicators | 16 | 0.830 | 851.6 | 1,752.4 |
| Harmonic + log-rainfall | 9 | 0.773 | 857.6 | 1,756.3 |

**Figure 4.**
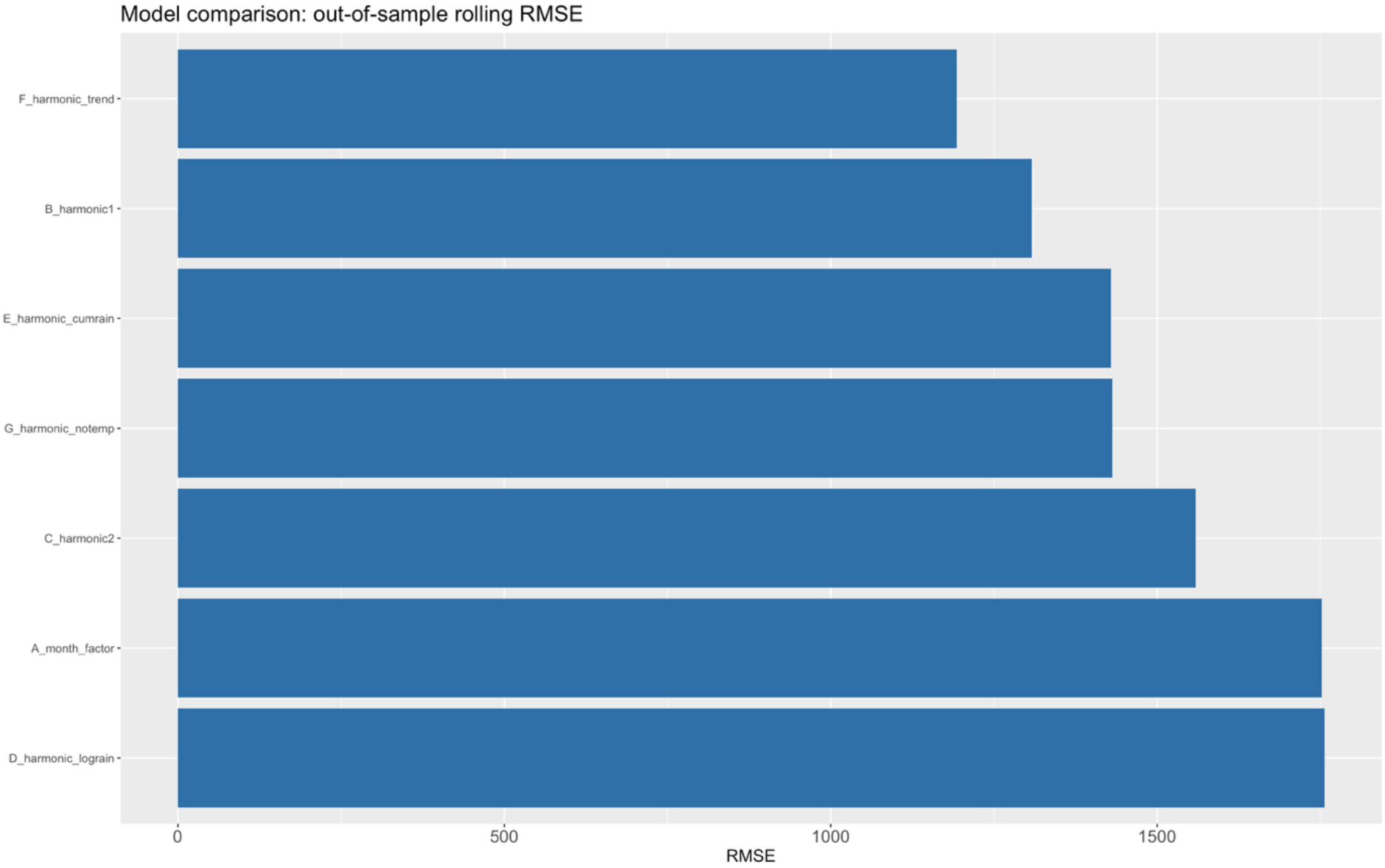
Comparison of the models.

### Climate predictors of dengue incidence

In the selected model, rainfall at both 1- and 2-month lag was significantly and positively associated with dengue incidence. A 1 mm increase in rainfall one month prior was associated with a 0.68% increase in expected monthly cases (IRR = 1.007, 95% CI: 1.003–1.011, *p* = 0.001), and a 1 mm increase two months prior with a 0.61% increase (IRR = 1.006, 95% CI: 1.002–1.011, *p* = 0.016). Temperature at 2- and 3-month lag showed no significant association with case counts (temp lag-2: IRR = 0.977, 95% CI: 0.903–1.056, *p* = 0.549; temp lag-3: IRR = 0.973, 95% CI: 0.909–1.041, *p* = 0.417). A significant negative linear time trend was also detected (β = −0.0093, *p* = 0.026), indicating a decline in monthly case counts over the study period independent of season and climate.

### Non-linearity of the rainfall effect

To assess whether the assumed log-linear rainfall effect was appropriate, a Negative Binomial generalized additive model (GAM) was fitted with penalized smooth terms for calendar month and each climate lag. The GAM explained a substantially larger share of deviance in-sample (89.2%) than the best linear GLM (83.3%; Table 1), driven primarily by non-linear rainfall terms: the effective degrees of freedom (edf) for rainfall at 1- and 2-month lag were 2.57 and 2.31, respectively (both *p* < 0.01), indicating genuine curvature rather than a linear dose-response relationship. Visual inspection of the fitted smooth terms showed that the rainfall effect was flat or slightly negative between 0 and approximately 80–100 mm, before rising sharply above this threshold — consistent with a minimum rainfall requirement for the accumulation of standing water needed for *Aedes* mosquito breeding. In contrast, the smooth terms for temperature at both lags collapsed to edf ≈ 1.0 with non-significant chi-square tests (*p* = 0.848 and *p* = 0.742), confirming the absence of a temperature effect found in the linear model.

Despite its superior in-sample fit, the GAM did not outperform the simpler harmonic GLM in out-of-sample rolling forecasts (RMSE = 1,273.0 vs. 1,192.9; Table 2), suggesting that the additional flexibility of the smooth terms, while capturing a real non-linear rainfall threshold, did not improve — and modestly reduced — forecasting performance given the limited sample size (n = 65 after lag construction).

**Figure 5.**
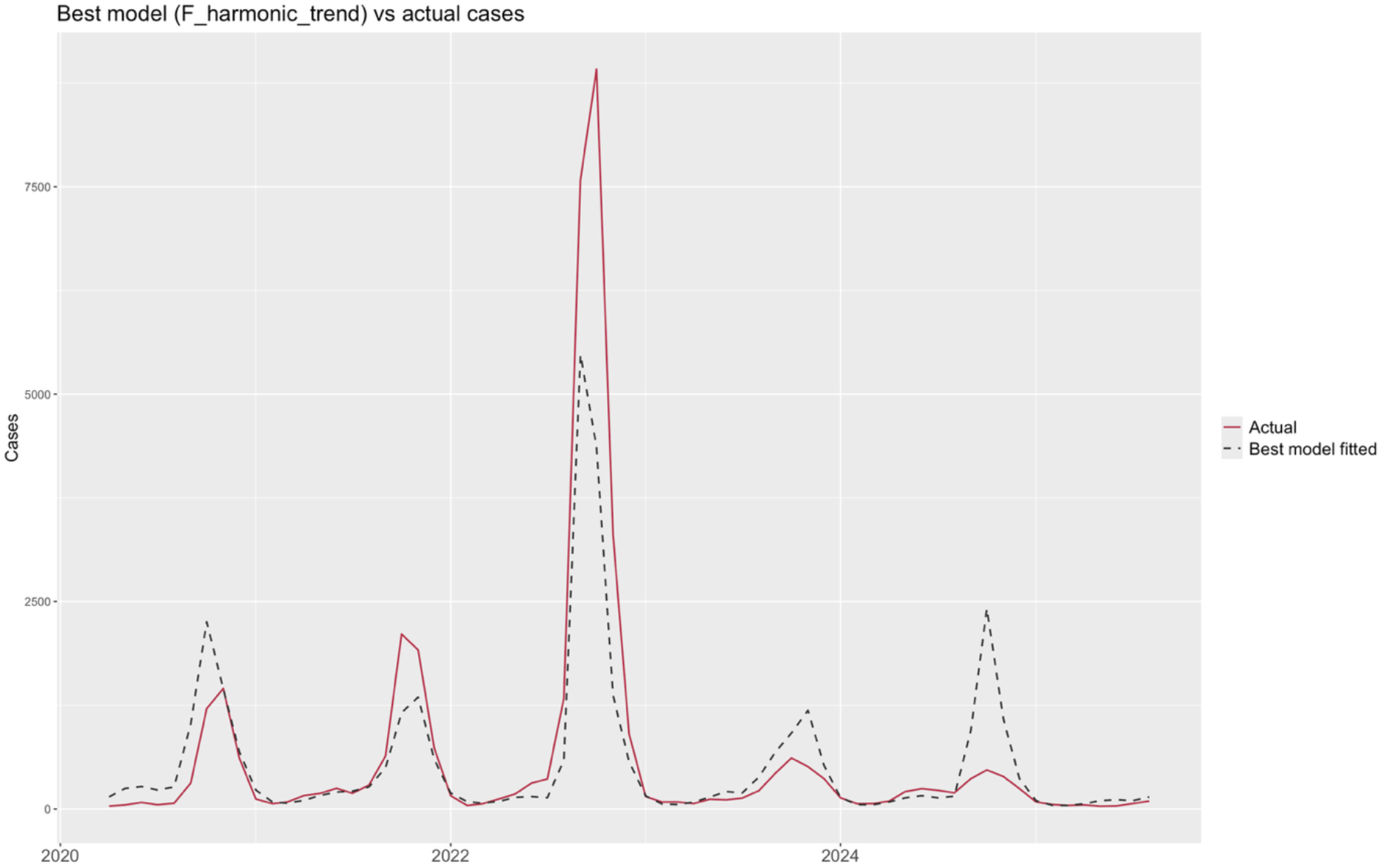
Fit of the best model (F Harmonic trend) to the actual dengue case data.

**Table 2.** Out-of-sample comparison of the best linear model and the GAM.

| **Model** | **Deviance explained (in-sample)** | **Out-of-sample RMSE** |
|---|---|---|
| Harmonic Negative Binomial GLM | 83.3% | **1,192.9** |
| Negative Binomial GAM | 89.2% | 1,273.0 |

Across all model specifications, rainfall at short lag (1–2 months) was a consistent, statistically significant predictor of dengue incidence, while temperature was not. The GAM analysis further indicated that this rainfall effect is non-linear, with an apparent threshold near 80–100 mm/month below which additional rainfall has little effect on case counts, and above which incidence rises sharply. A significant secular decline in cases was observed independent of climate drivers. These findings should be interpreted in light of the study’s limited temporal resolution (n = 65–68 monthly observations) and the dominance of a single extreme outbreak (September–October 2022) in the observed case series.

## Discussion

This study demonstrated the association between dengue incidence and meteorological factors, particularly rainfall and temperature, in Sindh, Pakistan between 2020 and 2025. The GAM fitted to the temperature data (Fig S1) adequately captures the clean annual temperature cycle with troughs (Jan/Feb) and peaks (Jul/Aug) recurring at nearly identical amplitude year over year until the end of 2025. The GAM fitted to the rainfall data also indicates the smoothed pattern of rainfall throughout the years. However, in July 2022, extreme rainfall occurred, causing disastrous impacts on health and infrastructure of the region [23].

Our study showed that rainfall at both 1- and 2-month lag was significantly and positively associated with dengue incidence. The results from our study corroborate the results from Bangladesh and Puerto Rico, both countries that have a high risk of dengue and observe recurrent monsoon rainfall [24-26]. This lag period corresponds well with the time from development of immature aquatic stages of the mosquito to the onset of human dengue cases at 27.1°C, the average daily mean temperature in Karachi between June-August [27]. These months observe high dengue case counts and frequent heavy rainfall events in our study area.

Similar to the findings in China, where a the greatest incidence rate ratio (IRR) of 1.37 [95% credible interval (CI): 1.02–1.83] occurred at a lag of 27 days, our analysis indicates an IRR = 1.007 [95% CI: 1.003–1.011] at a lag of 30 days and an IRR = 1.006 [95% CI: 1.002–1.011, p = 0.016] at a lag of 60 days [27].

The GAM analysis in our study showed a non-linear effect of rainfall on dengue virus cases, with an apparent threshold near 80–100 mm/month below which additional rainfall has little effect on case counts, and above which incidence rises sharply. The overall relationship between rainfall and dengue risk was non-linear, with a notable protective effect observed at moderate rainfall levels, particularly at short lags. This multifaceted role of rainfall has been noted in previous studies [28]. The risk during periods of very low rainfall or drought-like conditions may be linked to human behavior. During such times, residents often increase their reliance on domestic water storage containers, which can become stable and highly productive breeding sites for Aedes mosquitoes. Our findings, consistent with other research in Bangkok, suggest that moderate rainfall may provide a flushing effect on existing larval populations without creating widespread new breeding opportunities, thus explaining the observed protective effect [29].

In conclusion, the data show a significant impact of lagged rainfall on dengue virus case incidence, highlighting the importance of reactive flood control strategies to contain DENV case load in the country.

## Data Availability

All data produced in the present work are contained in the manuscript.

